# Automatic Electrophysiological Noise Reduction and Epileptic Seizure Detection for Stereoelectroencephalography

**DOI:** 10.1101/2021.06.16.21259055

**Authors:** Yufeng Zhou, Jing You, Fengjun Zhu, Anatol Bragin, Jerome Engel, Lin Li

## Abstract

The objective of this study was to develop a computational algorithm capable of locating artifacts and identifying epileptic seizures, which specifically implementing in clinical stereoelectroencephalography (SEEG) recordings. Based on the nonstationary nature and broadband features of SEEG signals, a comprehensive strategy combined with the complex wavelet transform (CWT) and multi-layer thresholding method was implemented for both noise reduction and seizure detection. The artifacts removal pipeline integrated edge artifact removal, discrete spectrum analysis, and peak density evaluation. For automatic seizure detection, integrated power analysis and multi-dynamic thresholding were applied. The F1-score was applied to evaluate overall performance of the algorithm. The algorithm was tested using expert-marked, double-blinded, clinical SEEG data from seven patients undergoing presurgical evaluation. This approach achieved the F1 score of 0.86 for noise reduction and 0.88 for seizure detection. This offline-approach method with minimum parameter tuning procedures and no prior information required, proved to be a feasible and solid solution for clinical SEEG data evaluation. Moreover, the algorithm can be improved with additional tuning and implemented with machine learning postprocessing pipelines.

## I. Introduction

Epilepsy is a chronic, noncommunicable neurological disorder that affects around 50 million people worldwide (World Health Organization, 2006) [1, 2]. A diagnosis of epilepsy requires the occurrence of at least one epileptic seizure as well as transient occurrence of signs and/or symptoms due to abnormal excessive or synchronous neuronal activity in the brain. [1]. Recurrent epileptic seizures might result in serious consequences, aggravating encephaledema, delaying the recovery of consciousness, and causing brain herniation, particularly in neurosurgery patients during the perioperative period [3]. Therefore, seizure control, especially recurrent seizure control, is an essential and critical step in epilepsy management, and it can be achieved through medication or surgical intervention. In most cases, surgical intervention by disconnecting epileptogenic brain regions is one of the most efficient therapies to treat recurrent refractory epilepsy [3,4,5,6].

Stereoelectroencephalography (SEEG) monitoring is required before surgical resection to precisely identify the seizure onset zone (SOZ). Review and interpretation of SEEG data play a critical role in guiding surgical resection [7, 8]. At present, it is widely accepted that the SEEG data and SOZ are manually reviewed and marked by a trained epileptologists in most epilepsy centers. While manual inspection of SEEG data can ensure a high precision rate, heavy labor costs and instability brought by variations in experience and ability among physicians cannot be ignored. A standardized SEEG analysis platform that minimizes the labor needed for data review and SOZ identification while also preserving diagnostic efficiency and accuracy is in demand.

The seizure event is the prominent electrophysiological pattern for SOZ identification. Digital signal processing (DSP) methods, such as time frequency approaches and dynamic thresholding methods, have been widely implemented in current clinical EEG reviews and feature identification routines. A solid technique with promising data processing speed and detection accuracy has been proved its effectiveness in previous study [9,10,11]. However, drawbacks of these approaches are either the lack of noise rejection capability that is inevitable during the recording, or the varied parameters that need to be adjusted when patients and the recording environment change.

Many recent seizure detection methods integrate machine learning, a majority of which apply supervised learning techniques to classify seizure and non-seizure events. Although some researchers have proven that Support vector machine (SVM) [12] and Random forest (RF) [13] have a high accuracy in a fixed training/test set, one downside of supervised learning is that the massive and precise labeling of seizure events is extremely time-consuming. Moreover, machine learning methods require that training and test sets originate from the same distribution, otherwise the test accuracy cannot be guaranteed. Many existing machine learning methods use online dataset for both training and testing, making then model inappropriate for real clinical dataset since the data distribution differs and the methods themselves lack preprocessing procedures. Consequently, a more adaptive and universal seizure detection technique with strong artifact removal capability is critical for clinical guidance.

In this project, a data processing pipeline that includes automatic noise reduction and seizure detection was established. The ultimate purpose of this pipeline was to largely reduce the labor costs for expert reviewing of clinical SEEG data. A primary aim was to provide a solution for efficiently and effectively labeling artifacts and seizure events while maintaining high precision and recall rates. In the noise cancellation section, a novel approach that accurately detected and removed artifacts was proposed. A fragmentation method [14] was adapted to the noise cancellation algorithm for computational efficiency. In the seizure detection section, a pipeline that combined fast spectrum power computation and multi-layer thresholding was established. Both these two sections did not require prior information and used universal parameters across all channels and patients.

## II. Methods

### A. Data collection

The SEEG data used in this project was collected using the Nihon Kohden EEG-1200 System for EEG monitoring in the Pediatric Epilepsy Center at the Shen Zhen Children’s Hospital. The Shen Zhen Children’s Hospital Research Ethics Committee approved this study, and informed consent was obtained from each patient. SEEG data was bandpass-filtered at 0.1–500.0 Hz and sampled at a rate of 2,000 Hz. Sampled data was quantified using a 16-bit analog-to-digital converter. SEEG data of each patient was obtained using 8-11 depth electrodes, with nine contacts per electrode and a contact surface of 0.8 mm^2^ per electrode. The electrodes on ECG, EOG, and EMG were excluded from this study. A total of seven patients who were subject to 12-14 hours of recording each were used for this study.

### B. Manual selection for noise and epileptic events

To fully estimate the SEEG data quality and set the gold standard to evaluate the algorithm, artifacts and epileptic seizure events were marked by two experts (J. You and F.J. Zhu). The data was first converted into European Data Format (EDF). A bipolar montage was applied to the recorded data to further eliminate global noise and achieve a clean view. EDF data was then loaded into the EDF browser and examined for overall quality. Data without a broken depth electrode and less than 50% of noise was selected for the event selection. To perform a manual event selection, a customized local field potential (LFP) signal reviewer was developed for the fast screening of raw SEEG data and marking of noise events and epileptic seizure activity. From the 98 hours of SEEG data from seven patients, a total of 1,637 noise events and 149 epileptic seizure events were selected for the algorithm evaluation.

### C. Automatic artifact detection

During the recording of the electrophysiological signals, various sources (i.e., external interferences, head and muscle motion, and abnormal electrical circuitry shortages) can contribute to artifact formation [15]. An efficient data preprocessing step is necessary to detect and remove artifacts before applying seizure detection procedures. Here, a novel approach that actively combines dynamic thresholding in frequency and time domains was proposed. The artifact reduction algorithm is composed of as the three following major steps.

#### Step 1: Edge artifacts removal

To achieve parallel computation and improve the processing efficiency, the entire time sequence was evenly split into multiple 2-minute epochs and therefore, as a result, edge artifacts were inevitably generated when involving Fourier transform computation in the following steps. A padding approach was introduced to resolve this problem. Specifically, the original signal was first flipped, then been concatenated on both sides of the original one. The extended sequence was then applied to computation. The advantage of applying the padding mechanism is that each timepoint was used the same number of times in convolution, which could successfully smooth the high power-level on edges.

#### Step 2:Discrete spectrum analysis

The clips extracted from step 1 were normalized and then used as input to obtain the spectrum power for further thresholding. Two time -frequency analysis methods were widely applied to compute power: the (1) Morlet wavelet transform and (2) Filter-Hilbert transform to overcome the nonstationary nature of SEEG. The Morlet wavelet transform was selected in this project for three main reasons. The first is that the Morlet wavelet always has a Gaussian shape in the frequency domain; thus, it is easier to obtain an ideal filter kernel compared to using the Filter-Hilbert method. The second reason is that the Morlet wavelet has a better computational efficiency in terms of obtaining the time frequency heat map. The third and most important reason is that the Filter-Hilbert method is only applicable in narrow band signals, and the wide band is essentially required for distinguishing artifacts from normal signals in noise cancellation.

Two critical variables were introduced to quantify and build up the characteristic energy thresholding to filter out artifacts after the normalized power *P*_*ij*_ was calculated, where *i* and *j* denoted the corresponding index of frequency and time vector, respectively. Specifically, the matrix *P* referred to a 2-min long time-frequency matrix and it was decomposed into multiple 1-second-long short clips for analysis. The first variable, the average power of the *k*^*th*^ clip across all frequency band 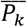 was denotes as (1).

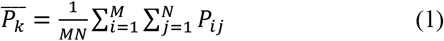

The term *M* and *N* represented the length of frequency vector and the number of points within the 1s window, respectively. The other variable *F*_*k*_, which referred to the high power-level coverage rate of the *k*^*th*^ clip. It was designed to check the number of frequencies that had average power greater than 0. Specifically, it was the denoted as (2).

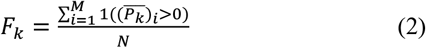

The term 1 was denoted as the function that retuned 1 if the condition inside the bracket was satisfied and returned 0 otherwise. The term 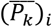 represented the mean value regraded to the time axis and it was defined as (3).

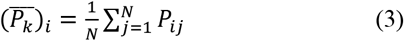

These two variables were used to compare with the average power threshold *P*_*thresh*_ (set to 2 in this project) and coverage rate threshold *F*_*thresh* (set to 0.75 in this project). Both these two thresholds were empirically selected. Only the clip that had 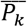 and *F*_*k*_ higher than *P*_*thresh* and *F*_*thresh* simultaneously would be considered as candidates of artifacts.

#### Step 3:Peak density evaluation

Previously steps were capable of filter out most common artifacts. However, frequency-based technique was hard to distinguish EEG spikes with large fluctuations and sharp transitions. Consequently, a peak density thresholding was integrated in the artifact detection pipeline. In this project, a moving window with a 1 second window size and a 0.2 second stride was implemented to quantify the fussy characters of the sequence. Number of peaks of the *k*^*th*^ clip was defined as followed: *PK*_*k*_ = [(*PK*_*k*_)_1_, (*PK*_*k*_)_2_, …, (*PK*_*k*_)_*n*_], which *n* denoted as the number of windows in the *k*^*th*^ clip. Specifically, the *i*^*th*^ element of vector *PK*_*k*_ was defined as (4).

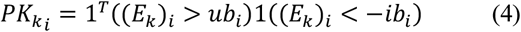

The term 1 here referred to the *N* dimensional vector that the entry equaled to 1 if the condition was satisfied, and 0 otherwise. (*E*_*k*_)_*i*_ denoted as the amplitude of the *i*^*th*^ slide of the *k*^*th*^clip. *N* represented the number of points in each slide. Terms *ub*_*i*_ and *lb*_*i*_ were defined as (5) and (6).

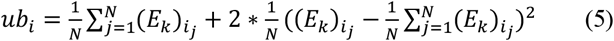

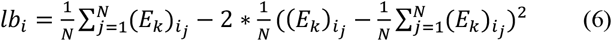

In this experiment, most EEG spikes were filtered out by setting the threshold to 3.5 times of the average number of peaks. If any sliding window had higher number of peaks than threshold, the epoch contained this certain window would be considered as candidate of artifact.

### D. Automatic seizure detection

Noise-free signals were served as the input of the seizure detection section. Two major steps were implemented in this section. The first is the integrated power (IP) quantification and thresholding and the second one is the multi-thresholding. The input sequence was split into multiple one-second-long clips and then, Morlet wavelet with the within the 3-30Hz frequency range was implemented to compute the spectrum power and the interpretation of selecting the range was based on the nature of electrophysiological seizures [16]. The IP in channel *i* and clip *j* was related with spectrum power *P*_*ij*_, and it was defined as (7)

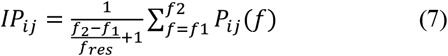

Noting that *f*1 and *f*2 were lower and upper bound of the frequency range, and *f*_*res*_ suggested the frequency resolution. In this experiment, they were empirically set to 3Hz, 18Hz, and 0.5HZ, respectively. *P*_*ij*_ (*f*) in (2) was different from the *P*_*ij*_ in (1) due to the following reasons: (i). They had different frequency range. (ii). Although, *P*_*ij*_ from (1) was also one-second-long, it was retrieved from a 2-min-long time-frequency matrix *P*, the *P*_*ij*_ in (2) was directly computed from the 1-second-long time window. *P*_*thresh*_ was introduced to filter out all clips that had low normalized power than it. The interpretation was that seizure events usually had higher amplitude than normal signals. In this project, *P*_*thresh*_ of the *i*^*th*^ channel was defined as (8).

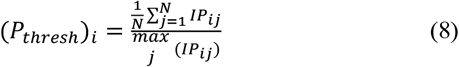

In (8), *N* referred to the total number of epochs in the *i*^*th*^channel.

The second step was the multi-thresholding. This part was composed of two steps, the time constrain and variance testing. A previous study proposed that the median lasting time of a human electrophysiological seizure should last at least 18.5 seconds [17], and hence, in this project the time constrain variable Δ*t* was set as 15 seconds which was slightly below the gold standard. The variables *t*_0_ and *t*_2_ denoted the first and the last consecutive clips that had the higher IP than the power threshold. Equation (9) defined the formula of time constrain variable.

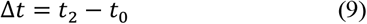

The purpose of the variance testing was to distinguish between SOZ and artifacts. The amplitude of seizure events in time domain would reflect a faster oscillation, but it was a significantly slower change comparing against artifacts. Consequently, the variance of the *i*^*th*^ channel and *j*^*th*^ epoch, *Var*_*ij*_ was implemented and it was defined as:

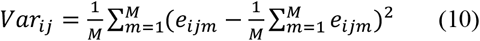

The term *M* denoted the number of points inside the epoch and *e*_*ijm*_ represented the voltage of the *m*^*th*^ point of the *ij*^*th*^ epoch. The averaged variance was implemented as the generic threshold. However, since the input sequence of this section has already passed through an artifact-removal pipeline, this project could skip the variance testing without compromising the performance of the detector.

### E. Statistical result

The distribution of positive and negative events was unbalanced in both sections, the accuracy was not optimal metric of evaluation. Confusion matrix was introduced to unbiasedly evaluate the performance of the algorithm. Four critical variables, true positives (TP), true negatives (TN), false positives (FP), and false negatives (FN) were applied to further quantify the matrix. TP was denoted as events which detector and reviewers labelled as artifacts/seizures. TN represented events that detector and reviewers determined events as normal signals. FP referred to events that algorithm marked as artifacts/seizures, but the decision was denied by experts. FN suggested that the detector misclassified true artifacts/seizures as normal ones. Then, F1-score (F1) along with precision rate (P) and recall rate (R) were calculated based on TP, TN, FP, and FN, and they were defined as:

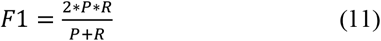

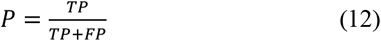

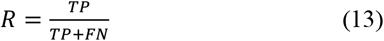

## III. Results

### A. Results of automatic noise reduction

Effectiveness of the automatic noise reduction algorithm has been reflected in Fig. 1. The raw plot of a two-minute clip of one selected channel was demonstrated in Fig. 1a. Spectrum representation of the noise signal from the local field potential revealed a wideband boosting in terms of power from 1 Hz to100 Hz (Fig. 1b). However, some specific neurophysiological patterns, such as K-complex (shown in the blue frame), were also misclassified as artifacts. To minimize the negative impacts brought by these patterns, a two-way thresholding approach, including characteristic energy thresholding (Fig. 1c) and peak density thresholding (Fig. 1d), was applied. Based on the observation, artifacts always had higher power-level, wider spanning characteristic energy pattern (Fig. 1c, red frame) compared to normal signals. The noise also revealed a rich peak density (Fig. 1d, red frame). Although the burst of power-level was also observed in the K-complex (Fig. 1b, blue frame) scenario, its’ duration was significantly squeezed in comparison with the true noise and more importantly, the energy (Fig. 1c, blue frame) and peak density (Fig. 1d, blue frame) were both substantially lower. After performing two dynamic thresholding procedures for characteristic energy and peak density, a total of 1,637 noise events were successfully detected.

**Figure 1.**
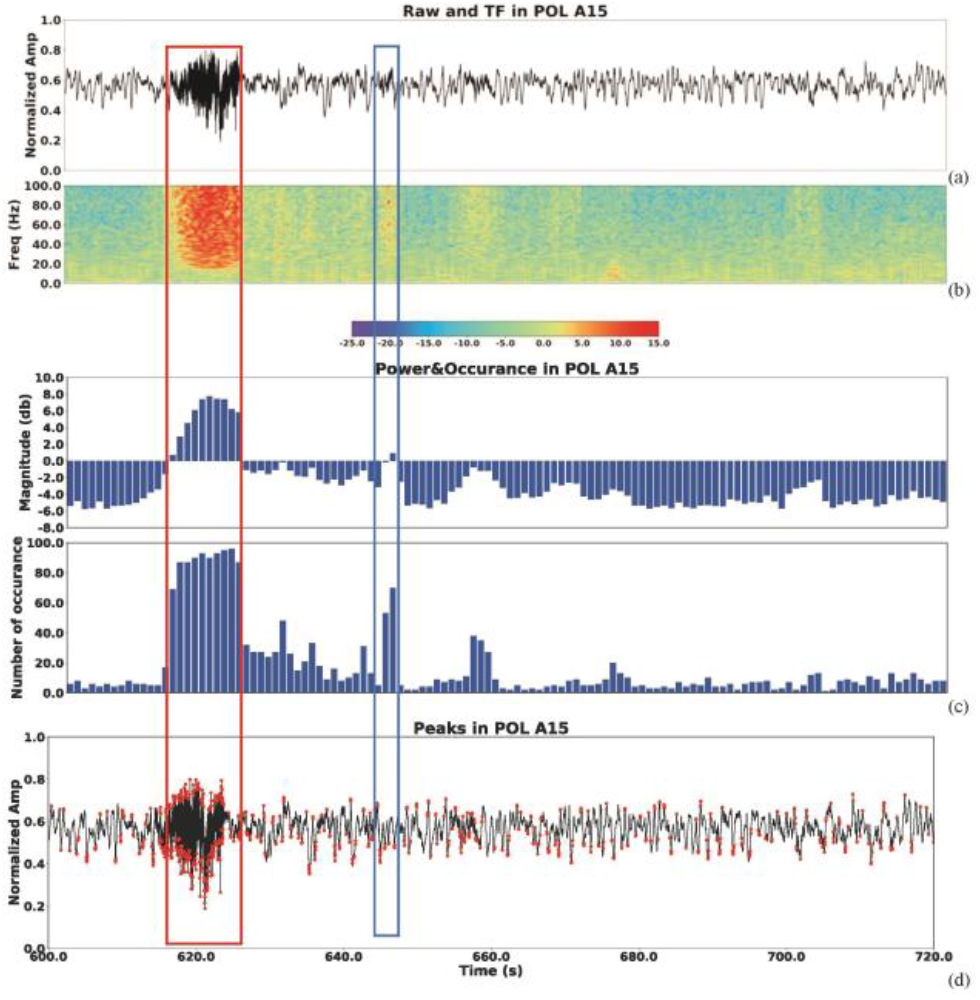
Illustration of the artifact removal procedures. (a): Raw data plot of SEEG signal with artifacts (red box). (b): Time-frequency analysis. (c): Power-level of each time bins (the 1^st^ row) and high power-level occurance of each time bin (the 2^nd^ row). (d): Peak detection of the ilustrated signals.

In the automatic noise reduction, six common brain areas with electrodes planted in were selected and each channel was treated separately to get its’ own F1, P, and R, which were calculated based on (11), (12), and (13). By comparing the testing results with the gold standard, the algorithm achieved a 0.868 precision rate in average, an overall 0.827 recall rate, and an 0.846 ± 0.031 F1 scores for all subjects across all channels (Table I). It was worth noting that the average F1 score was stable across most channels, except for PH16, which located in the CA1 area of the hippocampus.

**TABLE I.**
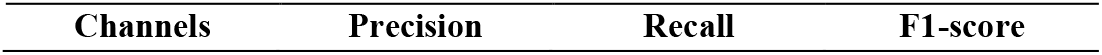
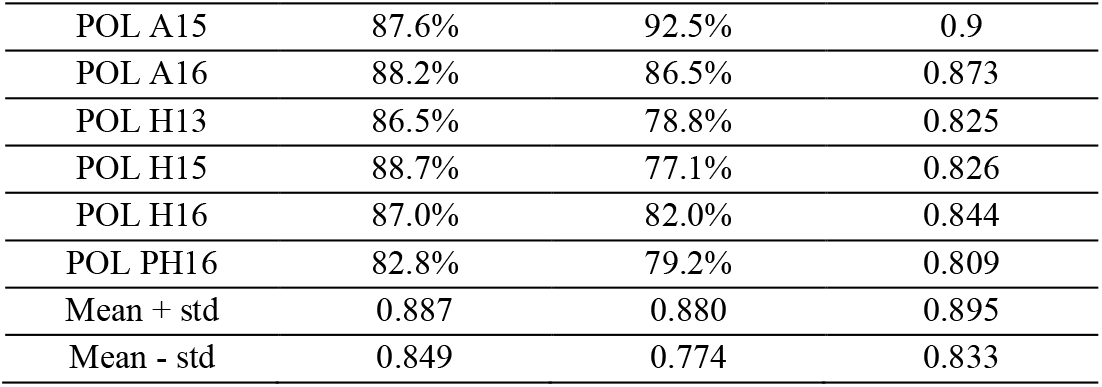
RESULT OF ARTIFACT REMOVAL ALGORITHM

| Channels | Precision | Recall | F1-score |
| --- | --- | --- | --- |
| POL A15 | 87.6% | 92.5% | 0.9 |
| POL A16 | 88.2% | 86.5% | 0.873 |
| POL H13 | 86.5% | 78.8% | 0.825 |
| POL H15 | 88.7% | 77.1% | 0.826 |
| POL H16 | 87.0% | 82.0% | 0.844 |
| POL PH16 | 82.8% | 79.2% | 0.809 |
| Mean + std | 0.887 | 0.880 | 0.895 |
| Mean - std | 0.849 | 0.774 | 0.833 |

### B. Results of automatic seizure reduction

The detailed processing procedures of automatic seizure detection was clearly demonstrated in Fig. 2. and Fig. 3. After IP thresholding, all possible candidates of seizure events were identified (Fig. 2a, red lines). In Fig. 3a, each bar represented a one-second clip, and the height of the bar represented the ratio between the integrated power and maximum integrated power of the whole sequence. The gray bars denoted time bins that were discarded by the IP threshold, and the blue bars suggested events that were kept after IP threshold since their relatively high power-level. The height of the red line in the *i*^*th*^ channel equaled to (*P*_*thresh*_)_*i*_ and it was defined in (8).

**Figure 2.**
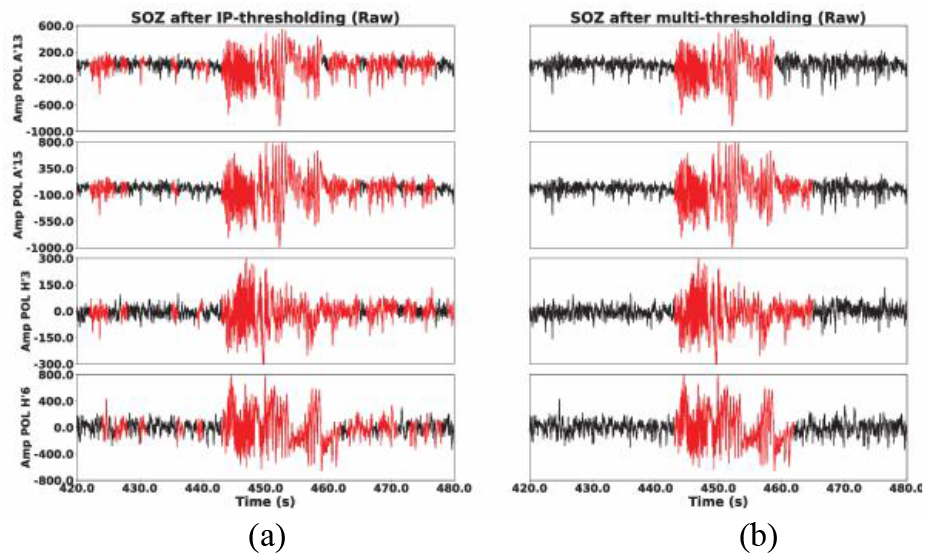
Plots of detected SOZ in a one-minute window. (a): Results that are only after IP thresholding. (b): Results that pass through all thresholding procedure.

**Figure 3.**
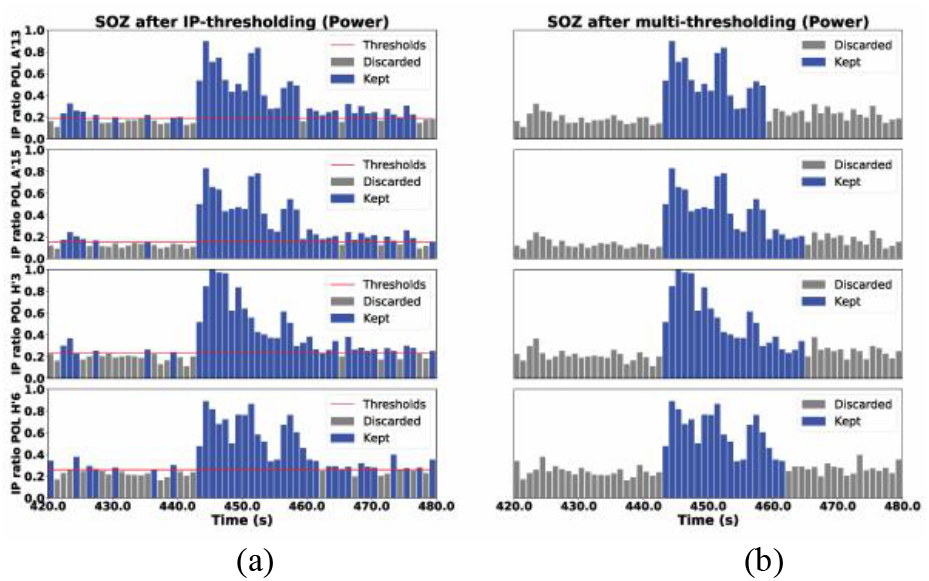
Illustration of IP in each time bin in a one-minute window. (a): Results with only IP thresholding analysis. (b): Results that pass through all thresholding procedures.

It was worth noting that a simple thresholding procedure based on the integrated power (Fig. 3a) was not sufficiently effective since epileptic spikes and slow wave spindles also had higher power than normal signals and thus, the multi-thresholding approach was further implemented to exclude the false positives. Unqualified epochs with shorter durations were removed (Fig. 2b, 3b).

The distribution of SOZ among all selected channels in a 50-minute recording were demonstrated in Fig. 4, suggesting that only time periods with high power-level could be considered as real seizure events (blue), while the rest of the events were marked as normal (gray). However, some high-power level events were also labelled as normal, and Fig. 5 demonstrated how these false positives were discarded by the following multi-thresholding procedure. The blue lines and gray lines denoted true positives and false positives of SOZ, respectively. Specifically, Fig. 5a and Fig. 5b informed that the duration time was the key reason that some seizure events with high power-level in Fig 4 were marked in gray rather than being considered as real seizure events.

**Figure 4.**
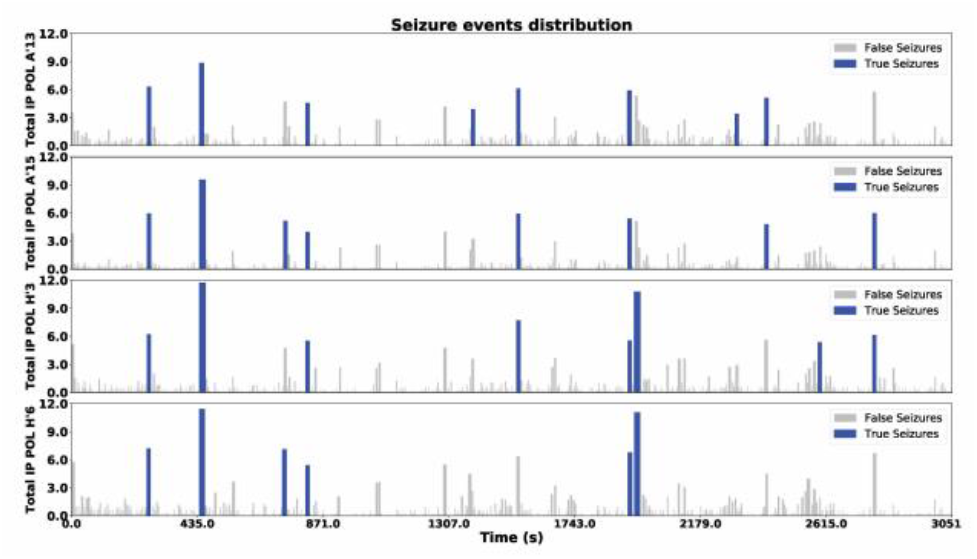
Seizure events distribution across all selected channels

**Figure 5.**
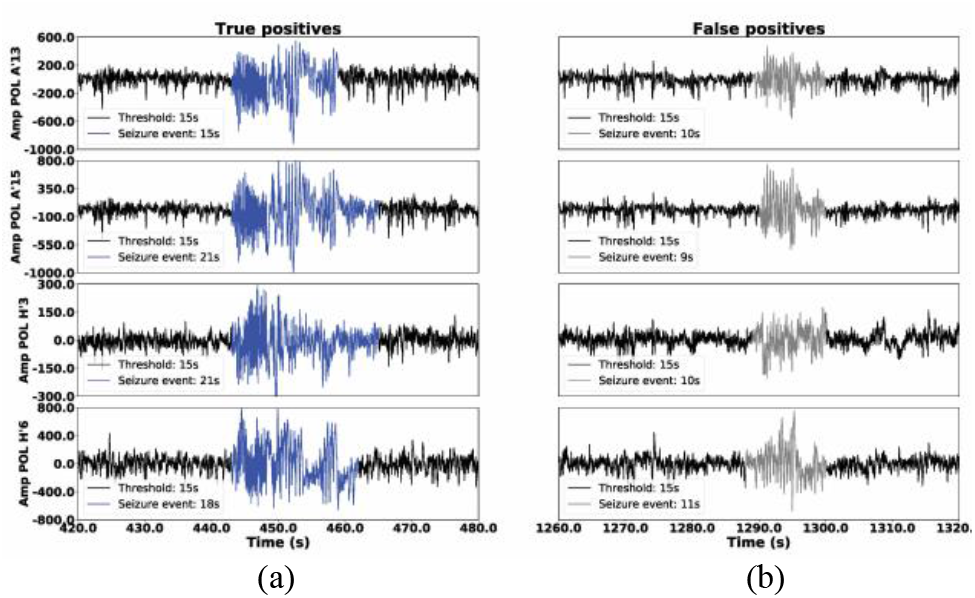
Demonstration of the effectiveness of time constrain analysis. (a): Events that are accepted by time-constrain analysis. (b): Events that are discarded by time-constrain analysis.

Unlike the noise reduction section which treated each channel separately, the seizure detection algorithm made the collective decision. For example, if a SOZ was detected in any one of the channels within the clip *i* and the result was confirmed by visual inspection, then the *i*^*th*^ clip would be marked as one TP. The other difference was that, in the seizure detection measurement, the length of the clip was set to 100 seconds rather than 1 previously. The detector has successfully detected 149 seizure events from roughly the 14 hours of recording. The processing time per run was within 3 minutes, excluding the figure plotting time, which was a decent speed considering that the Morlet wavelet was applied to the algorithm to compute the spectrum power. Table. II provided the confusion matrix of seizure detection algorithm, and therefore, the P, R and F1 could be calculated based on (5)-(7), and they were 87.6%, 87.6% and 0.876 respectively.

**TABLE II.** RESULT OF SEIZURE DETETOR

| Confusion matrix | True seizures | True normal |
| --- | --- | --- |
| Predicted seizures | 149(TP) | 21(FP) |
| Predicted normal | 21(FN) | 301 |

## IV. Conclusion

This study presented an efficient and accurate algorithm for automatic noise reduction and seizure detection specifically designed for SEEG measurements. These algorithms required no prior information regarding artifacts or seizures patterns, detecting characteristic changes in the frequency and amplitude of an entire SEEG recording by computing the power of each short clip via the Morlet wavelet. Specifically, dynamic thresholding procedures from both time and frequency domains were applied to exclude false positives. The parameters for thresholding were adapted for all SEEG channels.

Although the method proposed by Hopfengärtner [9] was more computationally efficient and had a solid performance in terms of detecting accuracy, one advantage of this method was that a solid noise reduction module being integrated into the entire pipeline, which in turn could be beneficial for reducing false positives in the seizure detection section. This approach implemented seizure detection in a channel-wise manner rather than combing all channels information and then taking the average. An advantage of this implementation was that channel-wise information could be served as a solid database to provide sufficient info to build up brain network to analysis the originality and patterns of seizure spread, which was critical as a clinical guidance. The seizure detection pipeline proposed by Fürbass et al. [10,11] had a strong noise reduction section, but it would consume more time in tuning thresholding parameters when applied to SEEG signals.

This algorithm could be improved in two ways in the future study. First, the preprocessing procedure in this project only concerned about noise. However, analyzing and classifying sleep stage data were additional essential preprocessing steps and have been overlooked this time. Fortunately, the frequency bands of sleep SEEG analysis and seizure detection highly overlap, and the multi-taper technique has been proven more effective than other time frequency analysis methods for processing sleep data [18, 19, 20]. As a result, in the future study, the multi-taper method could be considered to be embedded into the pipeline and replace the current Morlet-wavelet method after noise cancellation to simultaneously execute the computation workload for sleep data processing and seizure detection, which would make the algorithm more completed and robust without the loss of efficiency.

Second, as mentioned in the introduction, one weakness of a current machine learning-based method was the labor and time costs required for labeling. Saab et al. [21] proved that semi-supervised learning is applicable to automatic seizure detection. Consequently, the current approach could be used as a collector to automatically include high-quality true SOZ into the seizure library, then experts could select most representative ones from the database to serve as the training set for the semi-supervised learning technique, which could be beneficial for further improve the reliability of the detector.

## Data Availability

The authors confirm that the data supporting the findings of this study are available within the article or supplementary materials.

